# Factors associated with the uptake of Intermittent Preventive Treatment (IPTp-SP) for malaria in pregnancy: further analysis of the 2018 Nigeria Demographic and Health Survey

**DOI:** 10.1101/2022.06.23.22276819

**Authors:** Godwin Okeke Kalu, Joel M Francis, Latifat Ibisomi, Tobias Chirwa, Juliana Kagura

## Abstract

Pregnancy-associated malaria is preventable and curable with intermittent preventive treatment with Sulfodoxine-Pyrimethamine (IPTp-SP). However, despite the effectiveness of IPTp-SP against malaria in pregnancy, the uptake among pregnant women in Nigeria remains very low. Thus, this study aimed to establish the factors associated with the uptake of at least one dose and optimal doses of IPTp-SP among pregnant women aged 15 to 49 years living in Nigeria in 2018. The study included 12,742 women aged 15 to 49 years with live births two years before or during the 2018 Nigeria Demographic Health Survey (NDHS) in the analysis. Descriptive analysis was carried out to determine the prevalence of IPTp-SP uptake. Multivariable logistic regression was used to establish the factors associated with receiving IPTp-SP during pregnancy, adjusting for possible confounding factors. Given the complex survey design, all analyses adjusted for sampling weight, stratification and clustering. The p-value of <0.05 was considered significant. In 2018, the prevalence of at least one dose of IPTp-SP was 63.6% (95% CI:62.0–65.1), and optimal doses of IPTp-SP were 16.8% (95% CI:15.8–17.8) during pregnancy. After the multivariable analysis, age group, region, frequency of ANC visits, belief in IPTp-SP effectiveness, and morbidity caused by malaria predicted the uptake of at least one IPTp-SP dose. Similar maternal characteristics, including household wealth index, spouse’s educational level and media exposure, were significantly associated with taking optimal IPTp-SP doses. For instance, women in the wealthiest households whose husbands had secondary education predicted a four-fold increase in uptake of at least one IPTp-SP dose (aOR:4.17; 95% CI:1.11–8.85).

The low prevalence and regional variations of IPTp-SP uptake in the study area imply that most pregnant women in Nigeria are at substantial risk of pregnancy-associated malaria. Therefore, stakeholders should explore context-specific strategies to improve the IPTp-SP coverage across the regions in Nigeria.

## Introduction

Over the past two decades, about 1.7 billion malaria cases and 10.6 million malaria-related deaths were averted (1). Also, malaria mortality rates, that is, deaths per 100,000 population at risk, declined from 25 in 2000 to 12 in 2015 and 10 in 2019, globally (2). Equally, the estimated number of deaths decreased from 896,000 in 2000 to 558,000 in 2019 (2, 3). However, in 2020, malaria deaths increased to 627,000, partially due to disruptions of malaria services during the COVID-19 pandemic. In 2020, the WHO African region accounted for 95% of the 241 million cases and 94% of the global malaria deaths with four countries accounting for almost half of the global malaria deaths (2). These four countries are Nigeria (31.9%), the Democratic Republic of the Congo (13.2%), United Republic of Tanzania (4.1%), and Mozambique (3.8%) (2). Nonetheless, young children and pregnant women remain at significant risk of malaria infections. In 2019, 35% (12 million) of the estimated 33 million pregnant women living across 33 countries in the WHO African region were exposed to malaria infections during pregnancy (2). In the same year, the exposure to malaria infections during pregnancies resulted to 822,000 children born with low birthweight in these 33 countries (2)

In addition, approximately 50 million pregnant women remains at significant risk of pregnancy-associated malaria and possible malaria-related death annually (4). The risk associated with malaria in pregnancy can be drastically reduced by using Intermittent Preventive Treatment with Sulphadoxine-Pyrimethamine (IPTp-SP) for pregnancy-associated malaria as part of a multilevel three-staged approach (5). IPTp-SP utilisation among pregnant women has contributed to nearly 42% decrease in low birth weight, 38% decrease in neonatal death, and 65% decrease in placental malaria in SSA countries (1, 6). However, full benefits are evident when pregnant women receive three or more IPTp-SP doses (6). Receiving optimal IPTp-SP doses during pregnancy has increased the mean birth weight, reduced low birth weight, and fewer placental malaria than taking most two doses (6). The WHO revised IPTp-SP guidelines in 2012 to increase the number of Sulphadoxine-Pyrimethamine (SP) doses taken by pregnant women (5). Prior to 2012, the policy proposed that IPTp-SP should be given to pregnant women at every ANC visit to ensure they receive at least two SP doses whereas the revised policy recommends all pregnant women should receive one SP dose at every scheduled ANC visit except during first trimester at a one-month interval between each SP dose until time of delivery (5). Nigeria adopted the new IPTp-SP guideline in 2014 for preventing or treating malaria or asymptomatic malaria infection during pregnancy (7).

Despite the efforts by the WHO to increase the coverage of IPTp-SP in sub-Saharan African countries, several maternal characteristics have been established as a barrier to IPTp-SP uptake during pregnancy. Studies have reported that pregnant women’s attitudes and motivation to receive IPTp-SP was related to their levels of knowledge of malaria-related factors such as morbidities caused by malaria (8, 9). Similarly, region, religion, age, and marital status influenced the number IPTp-SP doses received during pregnancy (8,10,11). Socioeconomic considerations including education level, spouse’s education level, employment status, health insurance coverage and household wealth index influenced IPTp-SP uptake (12, 13). Increased uptake during pregnancy was associated with higher education (14, 15), and some studies found a strong association between being employed and IPTp-SP uptake (16). Also, several studies established that IPTp-SP uptake was pro-rich, such that women in the “middle” to “richest” household wealth index had higher IPTp-SP uptake compared to those in the “poorest” to “poorer” wealth categories, including uptake of at least three IPTp-SP doses during pregnancy (10,11,17,18).

The prevalence of at least one SP dose in Nigeria has increased from 27% in 2013 to 51% in 2015 and about 64% in 2018 (19). However, despite the relatively high prevalence of taking at least one dose of SP by pregnant women in Nigeria, the gap between uptake of the first dose of the antimalaria drug and at least three doses remains significantly large . For example, in 2018, the prevalence of at least one dose was 64% compared to the about 17% prevalence of three or more IPTp-SP doses (optimal doses) (19). This study examines the prevalence of IPTp-SP among pregnant women and factors associated with the uptake of at least one dose and three or more doses of IPTp-SP using the 2018 Nigeria Demographic Health Survey (NDHS).

## Methods

### Study design and study setting

The primary study; the Nigeria Demographic Health Survey women’s data, was a cross-sectional study conducted to provide estimates of the Nigerian populace’s basic demographic and health indicators. The survey included samples from 36 States, including the Federal Territory Capital (FCT). The sampling frame of the NDHS relied on the National Population and Housing Census (NPHC) carried out in 2006. Two sampling stages were carried out, and each of the 36 states and the Federal Capital Territory (FCT) were grouped into urban and rural areas to form 74 strata (19). First, 1389 census enumeration areas (EAs) were chosen from 74 strata using probability proportional to EA size as clusters independently. Secondly, 30 households were randomly selected without replacement from each cluster to form a sample size of 41,666 households. Weighting accurately represented the population during the NDHS dataset analysis due to the unequal distributed samples across each state and the potential disparity in the response rates. A total of 42,121 women, 15 to 49 years old, was scheduled for an interview, and 41,821 women were successfully interviewed (99% response rate). All women of childbearing age, between 15 to 49 years old, either resident in the selected household or female guests available in the households a night before 14 August to 29 December 2018, were eligible and interviewed. A detailed methodology for the 2018 NDHS has been published elsewhere (19).

### Study population

The study population were all women aged 15 to 49 years who had live births during or two years before the survey resident in Nigeria between 2016 and 2018. This study involved the secondary data analysis of the data obtained from the NDHS Women dataset collected between August to December 2018.

### Sampling and sample size

From the survey sampling frame, 41,821 women of childbearing ages (15 to 49 years) were interviewed. Among these women, 12,935 had livebirths during or two years preceding the 2018 NDHS. Of these, 12,742 women were extracted who reported to have received and known the exact number of IPTp-SP doses they took during their last pregnancy. These 12,742 women served as the analysis sample for this study after the survey weighting was applied (Fig 1).

**Fig 1:**
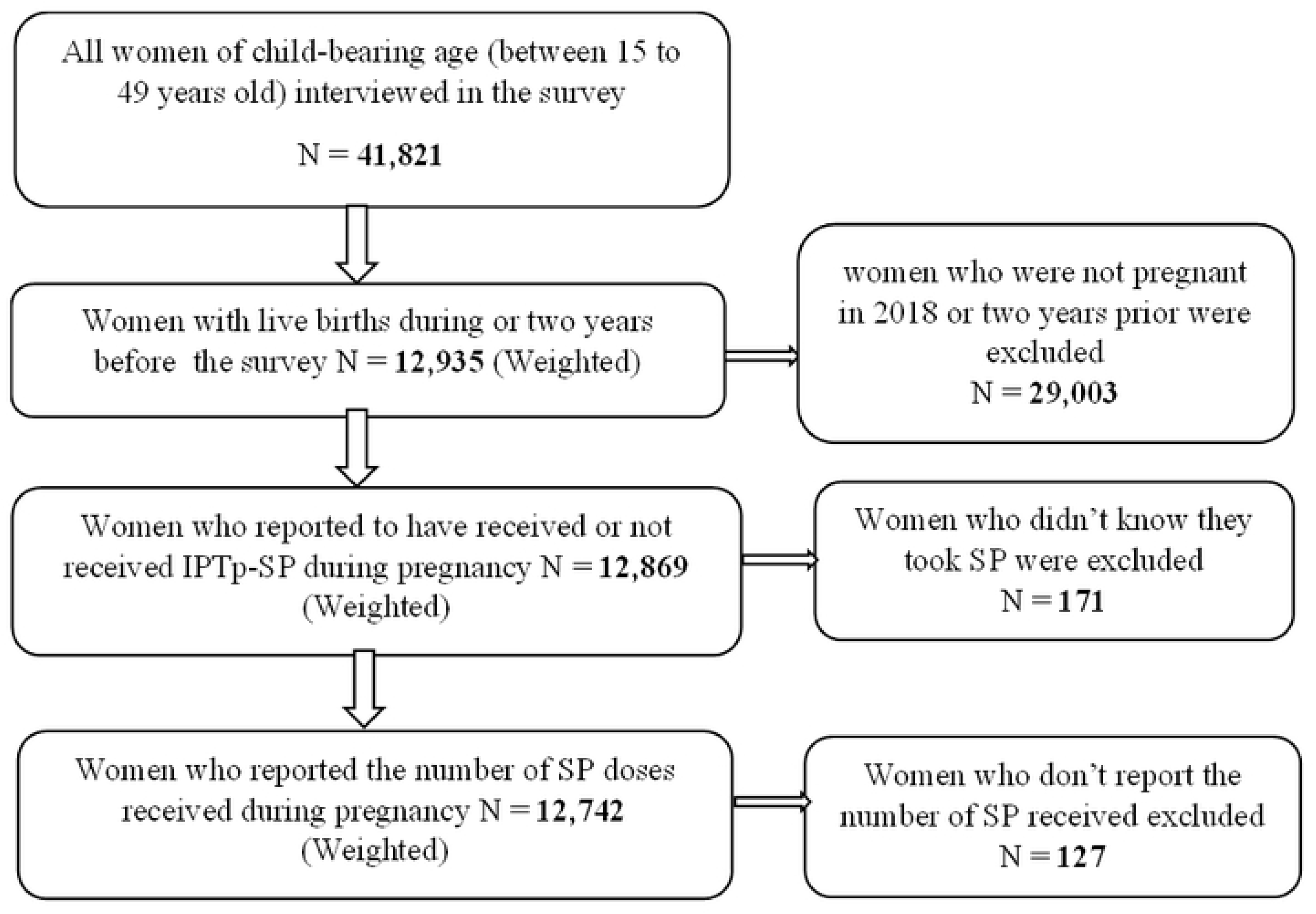
The Flow chart of the sample included in the NDHS 2018.

### Study variables

The extraction and coding of variables from the 2018 NDHS women dataset for the analysis were guided by previous literature (20, 21), and presented in Table 1.

**Table 1:** Categories and operational definitions of selected exposure variables.

| Exposure variables | Definitions/Categories |
| --- | --- |
| **Age in years** | Age of the respondent during the 2018 NDHS (15-24, 25-34, 35+) |
| **Marital status** | Unmarried (singles and formerly married; widows and divorced) and married, including co-habiting |
| **Household wealth Index** | Economic/wealth status of respondents' household (Poorest, poorer, middle class, richer, richest) |
| **Region or Geopolitical zone** | This is region or geopolitical zone where a respondent reside in Nigeria (North Central, North East, North West, South East, South South, South West) |
| **Residential areas** | This is the area of residence of respondents (urban, rural) |
| **Highest Educational Levels** | Respondent's formal level of education (such as; none, primary level, secondary level, higher level) |
| **Employment status** | This is respondent's employment status (Not employed, employed) |
| **Spouse's Education Level** | This is the formal education level of respondent's spouse or partner (none, primary, secondary, higher level) |
| **Frequency of ANC Visits** | This is the number times a respondent visited antenatal care (ANC) clinic during pregnancy ( None, less than 4 visits, 4+ visits) |
| **ANC Initiation** | The trimester a respondent commenced antimalaria treatment (IPTp-SP) during pregnancy (1st Trimester, 2nd trimester, 3rd trimester) |
| **Number of children born** | Total number children even born (Less than four children, four or more children) |
| **Belief in the effectiveness of IPTp-SP** | Respondent's belief or trust in the efficacy of IPTp_SP to protect them and their fetus against malaria infection during pregnancy (Low, Average, high) |
| **Belief about Malaria morbidity** | Respondent's belief or awareness of the morbidity caused by malaria in pregnancy (Low, Average, High) |
| **Health Insurance Coverage** | Respondent who subscribed to any form of health insurance (yes, no) |
| **Exposed to media messages** | This was generated from whether a respondent used any of newspaper/magazine, radio or television (yes, no) |

#### Outcome variables

The outcome variables are the uptake of at least one dose of IPTp-SP and optimal doses of IPTp-SP during pregnancy. The outcomes were created using responses from the following two NDHS survey questions (19): *“did you take SP/Fansidar to prevent you from getting malaria during pregnancy?* and *how many times did you receive SP/Fansidar during this pregnancy?”* For statistical analysis, the uptake of at least one dose of IPTp-SP was categorised and coded into 0 = no dose of IPTp-SP and 1 = at least one dose of IPTp-SP (≥1 dose). The uptake of optimal doses of IPTp-SP was categorised and coded into 0 = two or fewer doses (≤ 2 doses) and 1 = at least three doses of IPTp-SP (≥ 3 doses) (10, 21)

#### Exposure variables

The extracted exposure variables were sub-grouped into the following factors empirically or theoretically associated with uptake of IPTp-SP: sociodemographic, pregnancy-related, and knowledge of malaria-related factors. Sociodemographic factors include residential areas, age in years, household wealth index, the highest level of education, region, religion, employment and spouse’s educational attainment. Pregnancy-related factors include frequency of ANC visits, the timing of first ANC initiation, and parity. In addition, knowledge of malaria-related factors includes belief in the effectiveness of IPTp-SP, belief about malaria consequences, subscription to health insurance and media exposure. The household wealth index was derived in the primary study (NDHS 2018) by scoring each number and asset owned by the selected households using principal component analysis (PCA). The assets ranged from a television to a bicycle or car and housing characteristics such as the source of drinking water, toilet facilities, and flooring materials. Then, the National wealth quintiles or index compiled by allocating the household score to each usual (de jure) household resident. Subsequently, each 20% of the household population was divided into five equal categories from poorest to richest. (19).

The score for the level of belief in the effectiveness of IPTp-SP and malaria morbidity was generated using a three-point Likert scale (22). The level of belief score about the effectiveness of IPTp-SP from two survey questions (19): *“does malaria preventive medicine keep the mother healthy?” “Does malaria preventive medicine keep the baby healthy?”* Also, the level of belief score about malaria morbidity was derived from the following four survey questions (19): *“can malaria lead to death?” “can malaria make people dangerously sick?” “only weak children die of malaria?” “Do not worry about malaria can be cured?”* The women’s responses to the abovementioned questions asked in the Nigeria DHS were scored using a three-point Likert scale such that 1 = Disagree, 2 = Do not know, and 3 = Agree (22). After that, row mean scores were calculated for the women’s array of responses to these questions using the “egen” command on STATA. A score of one was the least possible score, and three was the highest possible score. Finally, the terciles of the composite scores were used as a cut-off to categorise the level of belief scores into low (1.0 – 1.9), average (2.0) and high scores (2.1 – 3.0).

### Data management and analysis

Data extraction, cleaning, re-coding and analysis were done using Stata version 16 (23). The extracted data were restricted to only women with live births during or two years before the 2018 NDHS. However, in creating the outcome variable, women who did not know if they took SP/Fansidar and the number of doses of SP/Fansidar taken was dropped, that is, 171 and 127 women, respectively. All selected exposure variables were categorical, and the missing observations across the exposure variables were reported as missing responses. Finally, the cleaned dataset was saved independently and converted into a survey dataset by applying the weighting, stratification and clustering. Proportions and frequencies were used to summarise all categorical variables for the descriptive analysis. At the same time, bivariate and multivariable logistic regression were used in the analytical analysis. The bivariate and multivariable logistic regression models were fitted using a four-step approach for modelling survey data as Heeringa et al., 2017 (59) and Hosmer and Lemeshow 2000 (60) recommended. The four-step approach is detailed below:

First, we fitted a bivariate logistic regression model to estimate the association of the outcome variables (at least one dose and optimal uptake) to each exposure variable. Second, all exposure variables with a p-value of <0.10 from the bivariate models were selected as candidates for main effects in the multivariable logistic regression model. Building the final multivariable logistic regression model involved an iterative process—the exposure variables of importance identified by previous studies were arranged from highest to the least importance. Third, fitted different model specifications by manually adding or dropping each exposure variable by performing the adjusted Wald test to assess each exposure variable’s contribution to the model. Lastly, scientifically plausible interactions among the exposure variables were checked. The interaction term was assessed by specifying factorial interaction using binary operators (##). The selection of the variables to interact was based on epidemiological notions or from previous literature. However, the selected multivariable logistic regression model accounted for only interaction terms with a p-value < 5%. Then, the fit of the final model was assessed using the Hosmer-Lemeshow goodness-of-fit test. The final models estimated the adjusted odds ratios (aORs). The significance level used was less than 5% (<0.05), two-tailed at 95% confidence intervals.

### Ethical consideration

The Nigeria DHS 2018 was conducted by the Nigerian Population Commission in collaboration with the National Malaria Elimination Programme (NMEP) of the Federal Ministry of Health, Nigeria. Before each interview during the 2018 NDHS survey, all respondents above 18 years provided written informed consent, and for those under 18 years of age written informed consent was obtained from the parent/guardian. The fieldworker ensured confidentiality throughout the survey process. The respondents’ records were coded and de-identified (19). For this study, permission for NDHS 2018 dataset for secondary data analysis has been obtained from ICF International – Measure DHS website. Ethics approval was obtained from the University of the Witwatersrand Human Research Ethics Committee (Medical) **– M2011103**.

## Results

### General characteristics of study participants

The study included 12,742 pregnant women between 15 and 49 years with live births from the 2018 NDHS. The mean age was 28.3 ± 6.7 years old. The study participants were selected from all the six regions (geo-political zones) in varying proportions. Of which majority of them were lived in Northwest Nigeria. 12,180 (95.6%) were married or living with a man, while 561 (4.4%) were single, divorced or widows. Of these women, 6692 (52.7%) had less than four children, whereas 6049 (47.5%) had four or more children. Also, more than half of the women, 7888 (61.9%), resided in rural Nigeria. In addition, the distribution of study participants varied depending on maternal characteristics, 5,766 (45.3%) of study participants had no formal education, and 4568 (35.9%) of them had husbands/spouses with no formal education. Although 8827 (69.3%) of the study participants were gainfully employed, only 2.0% subscribed to health insurance coverage. Additionally, 75.8% of them attended ANC services during pregnancy, of which 56.1% visited the ANC clinic more than four times in 2018. In the same year, almost half (47.1%) of the study participants initiated ANC services in their second trimester. Details of the characteristics of the study participants are presented in Table 2.

**Table 2:** Study characteristics of pregnant women aged 15 to 49 years old.

| Characteristics | Categories | Frequency N(%) |
| --- | --- | --- |
| **Total** |  | **12,742 (100%)** |
| **Sociodemographic characteristics** |  |  |
| **Age in years**<br>Mean age: 28.3 years old<br>Standard deviation: 6.7 years | 15-24 | 3842 (30.2) |
|  | 25-34 | 6242 (49.0) |
|  | 35+ | 2657 (20.9) |
| **Marital status** | Unmarried | 561 (4.4) |
|  | Married | 12,180 (95.6) |
| **Household wealth Index** | Poorest | 2,763 (21.7) |
|  | Poorer | 2,933 (23.0) |
|  | Middle | 2,636 (20.7) |
|  | Richer | 2,358 (18.5) |
|  | Richest | 2052 (16.1) |
| **Region** | North Central | 1,770 (13.9) |
|  | North East | 2,339 (18.4) |
|  | North West | 4,638 (36.4) |
|  | South East | 1,263 (9.9) |
|  | South-South | 1,126 (8.8) |
|  | South West | 1,606 (12.6) |
| **Residential areas** | urban | 4,854 (38.1) |
|  | rural | 7,888 (61.9) |
| **Highest Educational Levels** | No Education | 5,766 (45.3) |
|  | Primary | 1,856 (14.6) |
|  | Secondary | 4,050 (31.8) |
|  | Higher | 1,070 (8.4) |
| **Employment status** | Not Employed | 3,915 (30.7) |
|  | Employed | 8,827 (69.3) |
| **Spouse's Education Level** | No Education | 4,568 (35.9) |
|  | Primary | 1,624 (12.7) |
|  | Secondary | 4,116 (32.7) |
|  | Higher | 1,873 (14.7) |
|  | Missing | 561 (4.4) |
| **Pregnancy-related characteristics** |  |  |
| **Frequency of ANC Visits** | No ANC Visit | 3,081 (24.2) |
|  | < 4 ANC Visits | 2,350 (18.4) |
|  | ≥4 ANC Visits | 7,151 (56.1) |
|  | Missing Response | 160 (1.3) |
| **ANC Initiation** | 1st Trimester | 2,273 (17.8) |
|  | 2nd Trimester | 6,003 (47.1) |
|  | 3rd Trimester | 1,366 (10.7) |
|  | Missing | 3,100 (24.3) |
| **Number of children born** | Less than four children | 6692 (52.7) |
|  | Four or more children | 6049 (47.5) |

| Knowledge of malaria-related characteristics |  |  |
| --- | --- | --- |
| **Belief in Effectiveness of IPTp-SP** | Low | 200 (1.6) |
|  | Average | 504 (4.0) |
|  | High | 12,038 (94.5) |
| **Belief about Malaria morbidity** | Low | 1,614 (12.7) |
|  | Average | 3,869 (30.4) |
|  | High | 7,259 (57.0) |
| **Health Insurance Coverage** | No | 12,484 (98.0) |
|  | Yes | 258 (2.0) |
| **Exposed to media messages** | No | 11,823 (92.8) |
|  | Yes | 919 (7.2) |

#### Prevalence of IPTp-SP uptake among pregnant women in 2018

The estimated prevalence of at least one IPTp-SP dose was 63.6% (95% CI: 62.0–65.1), and three or more IPTp-SP doses were 16.8% (95% CI:15.9–17.8) among pregnant women in Nigeria (Table 3). Yet, the prevalence varied significantly depending on the maternal characteristics except for marital status. For instance, the prevalence of at least one SP was highest among women aged 25 to 34 years at 64.9% (95% CI: 63.0 – 66.7). Similarly, women aged 25 to 34 also reported the highest uptake of three or more SP doses at 17.7% (95% CI: 16.5 –19.0). The result indicated that the higher the educational attainment among women, the higher the number of IPTp-SP doses received during pregnancy. Also, among pregnant women, the number of IPTp-SP doses received during pregnancy increased depending on their spouse’s educational level. Likewise, there was an increasing prevalence of SP doses among pregnant women from the poorest to the wealthiest household wealth index (p<0.001). The result also established that the frequency of ANC visits was directly related to the number of SP doses they take during pregnancy (p <0.001). The prevalence of at least one IPTp-SP dose at 80.3% (95% CI: 78.8 – 81.7) as well as the prevalence of optimal SP doses at 23.3% (95% CI: 22.0 – 24.6) was the highest uptake was highest for those who attended more than four ANC visits during pregnancy. Also, early initiation of antenatal care was related to a higher prevalence of at least one dose and optimal doses of SP (p <0.001). The prevalence of taking at least one dose and optimal SP doses was significantly related to the belief in the effectiveness of IPTp-SP for protection against malaria in pregnancy (p<0.001). In addition, the prevalence of at least one IPTp-SP dose differed based on the level of belief about morbidities caused by malaria (p <0.001) but not the prevalence of taking optimal SP doses (p=0.242).

**Table 3.** : Prevalence of IPTp-SP dose received by pregnant women aged 15 to 49 years in 2018.

| Factors | Total N (%) | % Women who received $\geq 1$ dose | | p-value | % Women who received $\geq 3$ Doses | | p-value |
| --- | --- | --- | --- | --- | --- | --- | --- |
|  |  | n<sup>a</sup> (%) | 95% CI |  | n<sup>b</sup> (%) | 95% CI |  |
| **Uptake of SP dose** | **12742 (100%)** | **8098 (63.6)** | **62.0 – 65.1** |  | **2143 (16.8)** | **15.9 – 17.8** |  |
| **Age in years** |  |  |  | 0.015* |  |  | 0.010* |
| 15-24 | 3842 (30.2) | 2352 (61.2) | 58.8 – 63.6 |  | 578 (15.0) | 13.6 – 16.6 |  |
| 25-34 | 6242 (49.0) | 4049 (64.9) | 63.0 – 66.7 |  | 1103 (17.7) | 16.5 – 19.0 |  |
| 35+ | 2657 (20.9) | 1697 (63.9) | 61.3 – 66.7 |  | 462 (17.4) | 15.7 – 19.3 |  |
| **Region** |  |  |  | <0.001* |  |  | <0.001* |
| North Central | 1770 (13.9) | 1003 (56.7) | 53.2 – 60.1 |  | 264 (14.9) | 13.1 – 17.0 |  |
| North East | 2339 (18.4) | 1520 (65.0) | 61.8 – 68.0 |  | 329 (14.1) | 12.1 – 16.3 |  |
| North West | 4939 (36.4) | 2712 (58.5) | 55.1 – 61.7 |  | 503 (10.8) | 9.4 – 12.4 |  |
| South East | 1263 (9.9) | 1003 (79.5) | 76.4 – 82.2 |  | 493 (39.1) | 35.9 – 42.3 |  |
| South South | 1129 (8.8) | 837 (74.3) | 71.6 – 76.9 |  | 275 (24.4) | 21.6 – 27.6 |  |
| South West | 1606 (12.6) | 1023 (62.7) | 59.9 – 67.3 |  | 279 (17.4) | 14.6 – 20.5 |  |
| **Residential areas** |  |  |  | <0.001* |  |  | <0.001* |
| Urban | 4853 (38.1) | 3524 (72.6) | 70.4 – 74.7 |  | 1032 (21.3) | 19.6 – 23.0 |  |
| Rural | 7888 (61.9) | 4574 (58.0) | 55.9 – 60.1 |  | 1112 (14.1) | 13.0 – 15.3 |  |
| **Marital status** |  |  |  | 0.980 |  |  | 0.753 |
| Unmarried | 561 (4.4) | 357 (63.6) | 59.0 – 68.0 |  | 98 (17.4) | 14.1 – 21.3 |  |
| Married | 12,180 (95.6) | 7741 (63.6) | 61.9 – 65.2 |  | 2046 (16.8) | 15.8 – 17.9 |  |
| **Highest Educational level** |  |  |  | <0.001* |  |  | <0.001* |
| No Education | 5766 (45.3) | 2953 (51.2) | 48.7 – 53.7 |  | 597 (10.4) | 9.3 – 11.6 |  |
| Primary | 1856 (14.6) | 1259 (67.8) | 65.2 – 70.4 |  | 340 (18.3) | 16.4 – 20.4 |  |
| Secondary | 4050 (31.8) | 3008 (74.3) | 72.5 – 75.9 |  | 938 (23.2) | 21.6 – 24.9 |  |
| Higher | 1070 (8.4) | 878 (82.1) | 79.2 – 84.7 |  | 268 (25.0) | 21.7 – 28.8 |  |
| **Household Wealth Index** |  |  |  | <0.001* |  |  | <0.001* |
| Poorest | 2763 (21.7) | 1325 (47.9) | 44.8 – 51.1 |  | 335 (12.1) | 10.5 – 14.0 |  |
| Poorer | 2933 (23.0) | 1593 (54.3) | 51.2 – 57.4 |  | 323 (11.0) | 9.6 – 12.6 |  |
| Middle | 2636 (20.7) | 1793 (68.0) | 65.7 – 70.3 |  | 423 (16.1) | 14.3 – 18.0 |  |
| Richer | 2358 (18.5) | 1728 (73.3) | 70.7 – 75.7 |  | 550 (23.3) | 21.0 – 25.9 |  |
| Richest | 2052 (16.1) | 1659 (80.8) | 78.3 – 83.2 |  | 512 (24.9) | 22.3 – 27.8 |  |
| **Employment Status** |  |  |  | <0.001* |  |  | <0.001* |
| Not Employed | 3915 (30.7) | 2247 (57.4) | 54.8 – 60.0 |  | 539 (13.8) | 12.3 – 15.4 |  |
| Employed | 8827 (69.3) | 5851 (66.3) | 64.7 – 67.9 |  | 1604 (18.2) | 17.1 – 19.3 |  |
| **Spouse's Educational Level** |  |  |  | <0.001* |  |  | <0.001* |
| No Education | 4568 (35.9) | 2160 (47.3) | 44.6 – 47.0 |  | 392 (8.6) | 7.4 – 9.9 |  |
| Primary | 1624 (12.7) | 1058 (65.2) | 62.1 – 68.2 |  | 248 (15.3) | 13.3 – 17.5 |  |
| Secondary | 4116 (32.7) | 3020 (73.4) | 71.6 – 75.1 |  | 974 (23.7) | 22.0 – 25.4 |  |
| Higher | 1872 (14.7) | 1502 (80.3) | 77.9 – 82.5 |  | 431 (23.0) | 20.5 – 25.8 |  |
| Missing Response | 561 (4.4) |  |  |  |  |  |  |
| **Frequency of ANC Visits** |  |  |  | <0.001* |  |  | <0.001* |
| No ANC Visit | 3081 (24.1) | 508 (16.5) | 14.7 – 18.5 |  | 134 (4.4) | 3.4 – 5.5 |  |
| <4 ANC Visits | 2350 (18.4) | 1717 (73.1) | 70.7 – 75.3 |  | 297 (12.6) | 11.0 – 14.4 |  |
| ≥ 4 ANC Visits | 7151 (56.1) | 5739 (80.3) | 78.8 – 81.7 |  | 1667 (23.3) | 22.0 – 24.6 |  |
| Missing Response | 160 (1.3) |  |  |  |  |  |  |
| **Timing of ANC initiation** |  |  |  | <0.001* |  |  | <0.001* |
| 1st Trimester | 2273 (17.8) | 1813 (79.8) | 77.5 – 81.9 |  | 640 (28.2) | 25.8 – 30.6 |  |
| 2nd Trimester | 6003 (47.1) | 4752 (79.2) | 77.6 – 80.7 |  | 1177 (19.6) | 18.3 – 21.0 |  |
| 3rd Trimester | 1365 (10.7) | 1007 (73.8) | 70.9 – 76.5 |  | 186 (13.6) | 11.5 – 16.1 |  |
| Missing Response | 3100 (24.3) |  |  |  |  |  |  |
| **Number of children born** |  |  |  | <0.001* |  |  | <0.001* |
| Less than 4 children | 6692 (52.7) | 4438 (66.3) | 64.6 – 68.0 |  | 1226 (18.3) | 17.1 – 19.6 |  |
| Four or more children | 6049 (47.5) | 3660 (60.5) | 58.4 – 62.5 |  | 917 (15.2) | 14.0 – 16.4 |  |
| **Media Exposure** |  |  |  | <0.001* |  |  | <0.001* |
| No | 11822 (92.8) | 7349 (62.2) | 60.5 – 63.8 |  | 1855 (15.7) | 14.8 – 16.7 |  |
| Yes | 919 (7.2) | 749 (81.5) | 78.1 – 84.6 |  | 288 (31.3) | 26.9 – 36.2 |  |
| **Health Insurance coverage** |  |  |  | 0.001* |  |  | <0.001* |
| No | 12484 (98.0) | 7889 (63.2) | 61.6 – 64.8 |  | 2072 (16.6) | 15.6 – 17.6 |  |
| Yes | 257 (2.0) | 209 (81.2) | 71.6 – 88.0 |  | 72 (27.8) | 21.9 – 34.6 |  |
| **Belief in Effectiveness of SP** |  |  |  | <0.001* |  |  | <0.001* |
| Low level of belief | 200 (1.6) | 86 (43.0) | 35.7 – 50.7 |  | 14 (7.2) | 4.5 – 11.4 |  |
| Average level of belief | 504 (4.0) | 174 (34.4) | 28.6 – 40.7 |  | 52 (10.4) | 7.3 – 14.6 |  |
| High level of belief | 12038 (94.5) | 7838 (65.1) | 63.5 – 66.7 |  | 2077 (17.3) | 16.3 – 18.3 |  |
| **Belief about Malaria morbidity** |  |  |  | <0.001* |  |  | 0.242 |
| Low level of belief | 1614 (12.7) | 912 (56.5) | 52.4 – 59.2 |  | 240 (14.9) | 12.7 – 16.7 |  |
| Average level of belief | 3869 (30.4) | 5400 (62.8) | 61.2 – 65.9 |  | 648 (16.8) | 15.6 – 19.7 |  |
| High level of belief | 7258 (57.0) | 134 (65.5) | 63.4 – 67.2 |  | 1255 (17.3) | 15.8 – 18.2 |  |
| ANC: Antenatal Care,<br>*potential candidate for the multivariable logistic regression |  |  |  |  |  |  |  |

### Factors associated with uptake of at least one IPTp-SP dose

The results indicated that all selected maternal characteristics were independently associated with receiving at least one IPTp-SP dose during pregnancy (Table 4). Yet, after adjusting for other factors in the multivariable model, uptake of at least one IPTp-SP dose was significantly associated with pregnant women’s age group, the region where they live, and the frequency of ANC visits. Also, women’s belief in the effectiveness of SP for malaria treatment and about morbidities caused by malaria influenced their decision to initiate antimalaria treatment during pregnancy. For instance, pregnant women aged 35 and above had a 42% higher likelihood of receiving at least the first dose than those aged 15 to 24 (aOR:1.42; 95% CI: 1.12–1.81). Also, women living in the south-west were 50% less likely to receive at least one dose than North-Central (aOR: 0.50; 95% CI: 0.38 – 0.64). The results also indicated that those with four or more ANC visits had 61% greater odds of taking at least one SP dose than those with fewer ANC attendance (aOR: 1.61; 95% CI: 1.36–1.91). The timing of ANC initiation showed no significant association with using at least one IPTp-SP dose (p =0.138). Those who somewhat believed in the drug’s effectiveness were 50% less likely to receive at least a single SP dose than those with a minimal belief (aOR:0.50; 95% CI: 0.26–1.00). Conversely, the awareness of morbidities caused by malaria during pregnancy predicted a higher uptake of at least one SP dose (p =0.013). Those with firm beliefs were 33% more likely to receive at least a single IPTp-SP dose than those unaware of malaria-associated morbidities (aOR: 1.33; 95% CI: 1.10–1.62). The household wealth index and spouse’s educational level interacted significantly with the uptake of at least one SP dose (p=0.048). As a result, women in the wealthiest household whose husbands had secondary education had a four-fold higher odds of receiving at least one dose of IPTp-SP than those impoverished with no formal education (aOR:4.17; 95% CI:1.11–8.85).

**Table 4:** Factors associated with uptake of at least one SP dose among pregnant women.

| Factors | Total N (%) | Crude OR (95% CI)<br>Uptake of at least one dose | P-value | Adjusted OR (95%CI)<br>Uptake of at least one dose | P-value |
| --- | --- | --- | --- | --- | --- |
| **Socio-demographic Factors** |  |  |  |  |  |
| **Age in years** |  |  | **0.011<sup>a</sup>** |  | **0.008<sup>a</sup>** |
| 15-24 | 3,842 (30.2) | 1.00 |  | 1.00 |  |
| 25-34 | 6,242 (49.0) | 1.17 (1.06 - 1.29) | 0.003 | 1.10 (0.92 - 1.31) | 0.287 |
| 35+ | 2,657 (19.3) | 1.12 (0.98 - 1.28) | 0.100 | 1.42 (1.12 - 1.81) | 0.004 |
| **Region** |  |  | **<0.001<sup>a</sup>** |  | **<0.001<sup>a</sup>** |
| North Central | 1,770 (13.9) | 1.00 |  | 1.00 |  |
| North East | 2,339 (18.4) | 1.42 (1.17 - 1.73) | <0.001 | 2.05 (1.63 - 2.59) | <0.001 |
| North West | 4,639 (36.4) | 1.08 (0.88 - 1.31) | 0.465 | 2.53 (1.98 - 3.23) | <0.001 |
| South East | 1,263 (9.9) | 2.96 (2.36 - 3.71) | <0.001 | 1.33 (1.01 - 1.74) | 0.039 |
| South South | 1,126 (8.8) | 2.22 (1.817 - 2.700) | <0.001 | 1.61 (1.23 - 2.11) | 0.001 |
| South West | 1,606 (12.6) | 1.34 (1.08 - 1.66) | 0.007 | 0.50 (0.38 - 0.64) | <0.001 |
| **Residential areas** |  |  | **<0.001<sup>a</sup>** |  | **0.087<sup>a</sup>** |
| Urban | 4,854 (38.1) | 1.00 |  | 1.00 |  |
| Rural | 7,888 (61.9) | 0.52 (0.45 - 0.60) | <0.001 | 1.15 (0.98 - 1.36) | 0.087 |
| **Highest Educational Level** |  |  | **<0.001<sup>a</sup>** |  | **0.854<sup>a</sup>** |
| No Education | 5,766 (45.3) | 1.00 |  | 1.00 |  |
| Primary | 1,856 (14.6) | 2.01 (1.74 - 2.33) | <0.001 | 1.01 (0.84 - 1.22) | 0.902 |
| Secondary | 4,050 (31.8) | 2.75 (2.41 - 3.14) | <0.001 | 0.94 (0.77 - 1.15) | 0.530 |
| Higher | 1,070 (8.4) | 4.371 (3.53 - 5.42) | <0.001 | 0.92 (0.67 - 1.25) | 0.585 |
| **Household wealth Index** |  |  | **<0.001<sup>a</sup>** |  | **0.846<sup>a</sup>** |
| Poorest | 2,763 (21.7) | 1.00 |  | 1.00 |  |
| Poorer | 2,933 (23.0) | 1.29 (1.10 - 1.51) | 0.001 | 0.93 (0.71 - 1.21) | 0.583 |
| Middle | 2,636 (20.7) | 2.31 (1.97 - 2.72) | <0.001 | 1.07 (0.77 - 1.48) | 0.698 |
| Richer | 2,358 (18.5) | 2.98 (2.49 - 3.57) | <0.001 | 1.20 (0.69 - 2.08) | 0.515 |
| Richest | 2,052 (16.1) | 4.58 (3.75 - 5.61) | <0.001 | 0.94 (0.38 - 2.37) | 0.902 |
| **Employment Status** |  |  | **<0.001<sup>a</sup>** |  | **0.693<sup>a</sup>** |
| Not Employed | 3,915 (30.7) | 1.00 |  | 1 |  |
| Employed | 8,827 (69.3) | 1.46 (1.31 - 1.62) | <0.001 | 1.03 (0.89 - 1.19) | 0.693 |
| **Spouse's Education Level** |  |  | **<0.001<sup>a</sup>** |  | **0.695<sup>a</sup>** |
| No Education | 4,568 (35.6) | 1.00 |  | 1.00 |  |
| Primary | 1,624 (12.7) | 2.09 (1.77 - 2.46) | <0.001 | 1.10 (0.66 - 1.82) | 0.721 |
| Secondary | 4,116 (32.7) | 3.07 (2.69 - 3.51) | <0.001 | 1.04 (0.70 - 1.51) | 0.882 |
| Higher | 1,872 (14.7) | 4.54 (3.80 - 5.42) | <0.001 | 1.95 (0.61 - 6.36) | 0.261 |
| **Pregnancy-related factors** |  |  |  |  |  |
| **Frequency of ANC visits** |  |  | **<0.001<sup>a</sup>** |  | **<0.001<sup>a</sup>** |
| <4 ANC Visits | 2,351 (24.7) | 1.00 |  | 1.00 |  |
| ≥ 4 ANC Visits | 7,151 (75.3) | 1.50 (1.31 - 1.72) | <0.001 | 1.61 (1.36 - 1.91) | <0.001 |
| **Timing of ANC Initiation** |  |  | **0.001<sup>a</sup>** |  | **0.138<sup>a</sup>** |
| 1st Trimester | 2,273 (23.6) | 1.00 |  | 1.00 |  |
| 2nd Trimester | 6,003 (62.3) | 0.96 (0.83 - 1.12) | 0.631 | 0.98 (0.83 - 1.15) | 0.780 |
| 3rd Trimester | 1,365 (14.2) | 0.71 (0.59 - 0.86) | <0.001 | 0.81 (0.65 - 1.02) | 0.080 |
| **Number of children born** |  |  | **<0.001<sup>a</sup>** |  | **0.328<sup>a</sup>** |
| <4 children | 6692 (52.7) | 1.00 |  | 1.00 |  |
| 4+ children | 6049 (47.5) | 0.78 (0.71 – 0.85) | <0.001 | 0.92 (0.77 – 1.09) | 0.328 |
| **Knowledge of malaria-related factors** |  |  |  |  |  |
| **Media Exposure** |  |  | **<0.001<sup>a</sup>** |  | **0.175<sup>a</sup>** |
| No | 11,823 (92.8) | 1.00 |  | 1.00 |  |
| Yes | 919 (7.2) | 2.69 (2.14 - 3.37) | <0.001 | 1.20 (0.92 - 1.56) | 0.175 |
| **Health Insurance coverage** |  |  | **0.001<sup>a</sup>** |  | **0.082<sup>a</sup>** |
| No | 12,484 (98.0) | 1.00 |  | 1.00 |  |
| Yes | 258 (2.0) | 2.51 (1.47 - 4.28) | 0.001 | 1.51 (0.26 - 1.00) | 0.053 |
| **Belief in Effectiveness of SP** |  |  | **<0.001<sup>a</sup>** |  | **<0.001<sup>a</sup>** |
| Low belief | 200 (1.6) | 1.00 |  | 1.00 |  |
| Average belief | 504 (4.0) | 0.69 (0.47 - 1.03) | 0.072 | 0.50 (0.26 – 1.00) | 0.053 |
| High belief | 12,038 (94.5) | 2.47 (1.81 - 3.38) | <0.001 | 1.51 (0.90 – 2.51) | 0.116 |
| **Belief about Malaria morbidity** |  |  | **<0.001<sup>a</sup>** |  | **0.013<sup>a</sup>** |
| Low belief | 1,615 (12.7) | 1.00 |  | 1.00 |  |
| Average belief | 3,869 (30.4) | 1.38 (1.17 - 1.62) | <0.001 | 1.32 (1.05 - 1.67) | 0.017 |
| High belief | 7,258 (57.0) | 1.49 (1.29 - 1.72) | <0.001 | 1.33 (1.10 – 1.62) | 0.003 |
| *ANC – Antenatal care*<br><sup>a</sup>*p-values – overall p-values for each exposure variable in the model, CI – Confidence Intervals*<br>*OR – Odds ratios, crude OR (from Bivariate analysis) Adjusted OR (from Multivariable analysis)*<br>**Goodness-of-fit of the model = $F(9, 1262) = 0.651$; $p = 0.754$**<br>**1 => Reference category** |  |  |  |  |  |

### Factors associated with the uptake of optimal doses of IPTp-SP

All included maternal characteristics in the study were independently associated with the uptake of at least three IPTp-SP doses (Table 5). However, after adjusting for all factors in the model, household wealth index, frequency of ANC visits, and ANC initiation were significantly related to taking at least three IPTp-SP doses. The result established that compared to women with no formal education, those with higher education were 30% less likely to take at least three SP doses (aOR: 0.70 95% CI:0.51–0.98). In contrast, women whose spouses attained higher education had an 85% probability of completing optimal doses than those whose spouses had no formal education (aOR: 1.85; 95% CI: 1.39–2.46). Unexpectedly, the uptake of optimal IPTp-SP doses was inversely associated with the household wealth index. Compared to women in most impoverished households, the probability of taking at least three SP doses was reduced by 35% among women in poorer households and middle class (aOR: 0.65; 95% CI:0.52–0.82) and decreased by 29% among wealthiest households (aOR: 0.71; 95% CI: 0.52 – 0.97). Furthermore, the probability of completing at least three doses among pregnant women who attended four or more ANC visits was 57% higher than those with fewer ANC visits (aOR: 1.57; 95% CI:1.31 – 1.88). Conversely, the timing of initiating ANC services predicted a negative association with taking optimal SP doses during pregnancy (p <0.001). Those who initiated ANC in their third trimester were 39% less likely to complete optimal IPTp-SP doses during pregnancy (aOR:0.61; 95% CI: 0.47-0.79). Also, pregnant women exposed to media messages were 1.39 times as likely to complete optimal doses of IPTp-SP than those not exposed to media messages (aOR: 1.39; 95% CI: 1.10 – 1.75). Also, the receipt of optimal doses was associated with the level of belief about morbidity caused by malaria (p = 0.021). In contrast, the belief level in the effectiveness of SP against malaria in pregnancy did not play a role in taking IPTp-SP (p=0.102)

**Table 5.** : Factors associated with uptake of optimal doses of IPTp-SP among pregnant women.

| Factors | Total N (%) | Crude OR (95% CI)<br>Uptake of optimal doses | p-value | Adjusted OR (95% CI)<br>Uptake of optimal doses | p-value |
| --- | --- | --- | --- | --- | --- |
| **Sociodemographic factors** |  |  |  |  |  |
| **Age in years** |  |  | **0.015<sup>a</sup>** |  | **0.583<sup>a</sup>** |
| 15-24 | 3,842 (30.2) | 1.00 |  | 1.00 |  |
| 25-34 | 6,242 (49.0) | 1.21 (1.06 - 1.38) | 0.004 | 1.07 (0.90 - 1.27) | 0.420 |
| 35+ | 2,462 (19.3) | 1.19 (1.01 - 1.40) | 0.036 | 1.14 (0.89 - 1.46) | 0.300 |
| **Region** |  |  | **<0.001<sup>a</sup>** |  | **<0.001<sup>a</sup>** |
| North Central | 1,770 (13.9) | 1.00 |  | 1.00 |  |
| North East | 2,339 (18.4) | 0.93 (0.74 - 1.18) | 0.558 | 0.90 (0.70 - 1.15) | 0.116 |
| North West | 4,639 (36.4) | 0.69 (0.56 - 0.86) | 0.001 | 0.97 (0.76 - 1.24) | 0.475 |
| South East | 1,263 (9.9) | 3.65 (2.98 - 4.48) | <0.001 | 2.96 (2.31 - 3.80) | 0.187 |
| South South | 1,126 (8.8) | 1.84 (1.48 - 2.30) | <0.001 | 1.58 (1.20 - 2.06) | <0.001 |
| South West | 1,606 (12.6) | 1.20 (0.28 - 1.55) | 0.166 | 0.82 (0.63 - 1.06) | <0.001 |
| **Residential areas** |  |  | **<0.001<sup>a</sup>** |  | **0.240<sup>a</sup>** |
| Urban | 4,854 (38.1) | 1.00 |  | 1.00 |  |
| Rural | 7,888 (61.9) | 0.61 (0.53 - 0.70) | <0.001 | 1.11 (0.93 - 1.31) | 0.240 |
| **Highest Educational Level** |  |  | **<0.001<sup>a</sup>** |  | **0.168<sup>a</sup>** |
| No Education | 5,766 (45.3) | 1.00 |  | 1.00 |  |
| Primary | 1,856 (14.6) | 1.94 (1.63 - 2.31) | <0.001 | 0.97 (0.80 - 1.17) | 0.749 |
| Secondary | 4,050 (31.8) | 2.61 (2.25 - 3.04) | <0.001 | 0.85 (0.68 - 1.03) | 0.115 |
| Higher | 1,070 (8.4) | 2.89 (2.31 - 3.62) | <0.001 | 0.70 (0.51 - 0.98) | 0.035 |
| **Household Wealth Index** |  |  | **<0.001<sup>a</sup>** |  | **0.003<sup>a</sup>** |
| Poorest | 2,763 (21.7) | 1.00 |  | 1.00 |  |
| Poorer | 2,933 (23.0) | 0.90 (0.72 - 1.11) | 0.326 | 0.65 (0.52 - 0.82) | <0.001 |
| Middle | 2,636 (20.7) | 1.38 (1.12 - 1.71) | 0.003 | 0.65 (0.51 - 0.82) | <0.001 |
| Richer | 2,358 (18.5) | 2.20 (1.78 - 2.73) | <0.001 | 0.82 (0.65 - 1.04) | 0.088 |
| Richest | 2,052 (16.1) | 2.41 (1.93 - 2.30) | <0.001 | 0.71 (0.52 - 0.97) | 0.027 |
| **Employment Status** |  |  | **<0.001<sup>a</sup>** |  | **0.668<sup>a</sup>** |
| Not Employed | 3,915 (30.7) | 1.00 |  | 1.00 |  |
| Employed | 8,827 (69.3) | 1.39 (1.21 - 1.60) | <0.001 | 0.97 (0.82 - 1.13) | 0.668 |
| **Spouse's Educational Level** |  |  | **<0.001<sup>a</sup>** |  | **<0.001<sup>a</sup>** |
| No Education | 4,568 (35.6) | 1.00 |  | 1.00 |  |
| Primary | 1,624 (12.7) | 1.92 (1.55 - 2.39) | <0.001 | 1.06 (0.84 - 1.34) | 0.611 |
| Secondary | 4,116 (32.7) | 3.30 (2.76 - 3.93) | <0.001 | 1.70 (1.39 - 2.08) | <0.001 |
| Higher | 1,872 (14.7) | 3.19 (2.568 - 3.961) | <0.001 | 1.85 (1.39 - 2.46) | <0.001 |
| **Pregnancy-related factors** |  |  |  |  |  |
| **Frequency of ANC Visits** |  |  | **<0.001<sup>a</sup>** |  | **<0.001<sup>a</sup>** |
| <4 ANC Visits | 2,351 (24.7) | 1.00 |  | 1.00 |  |
| ≥4 ANC Visits | 7,151 (75.3) | 2.10 (1.79 - 2.47) | <0.001 | 1.57 (1.31 - 1.88) | <0.001 |
| **Timing of ANC Initiation** |  |  | **<0.001<sup>a</sup>** |  | **<0.001<sup>a</sup>** |
| 1st Trimester | 2,273 (23.6) | 1.00 |  | 1.00 |  |
| 2nd Trimester | 6,003 (62.3) | 0.62 (0.54 - 0.71) | <0.001 | 0.71 (0.61 - 0.82) | <0.001 |
| 3rd Trimester | 1,365 (14.2) | 0.40 (0.32 - 0.50) | <0.001 | 0.61 (0.47 - 0.79) | <0.001 |
| **Number of children born** |  |  | **<0.001<sup>a</sup>** |  | **0.681<sup>a</sup>** |
| <4 children | 6692 (52.7) | 1.00 |  | 1.00 |  |
| 4+ children | 6049 (47.5) | 0.80 (0.72 - 0.89) | <0.001 | 0.96 (0.81 - 1.14) | 0.681 |
| **Knowledge of malaria-related factors** |  |  |  |  |  |
| **Media Exposure** |  |  | **<0.001<sup>a</sup>** |  | **0.007<sup>a</sup>** |
| No | 11,823 (92.8) | 1.00 |  | 1.00 |  |
| Yes | 919 (7.2) | 2.45 (1.96 - 3.07) | <0.001 | 1.38 (1.10 - 1.75) | 0.007 |
| **Health Insurance coverage** |  |  | **<0.001<sup>a</sup>** |  | **0.076<sup>a</sup>** |
| No | 12,484 (98.0) | 1.00 |  | 1.00 |  |
| Yes | 258 (2.0) | 1.94 (1.41 - 2.66) | <0.001 | 1.37 (0.97 - 1.94) | 0.076 |
| **Belief in Effectiveness of SP** |  |  | **<0.001<sup>a</sup>** |  | **0.102<sup>a</sup>** |
| Low | 200 (1.6) | 1.00 |  | 1.00 |  |
| Average | 504 (4.0) | 1.49 (0.78 - 2.85) | 0.223 | 1.17 (0.54 - 2.55) | 0.694 |
| High | 12,038 (94.5) | 2.68 (1.62 - 4.44) | <0.001 | 1.63 (0.92 - 2.88) | 0.093 |
| **Belief about Malaria morbidity** |  |  | **0.074<sup>a</sup>** |  | **0.021<sup>a</sup>** |
| Low | 1,615 (12.7) | 1.00 |  | 1.00 |  |
| Average | 3,869 (30.4) | 1.25 (1.01 - 1.54) | 0.037 | 1.14 (0.91 - 1.42) | 0.250 |
| High | 7,258 (57.0) | 1.20 (1.01 - 1.43) | 0.041 | 0.90 (0.73 - 1.10) | 0.305 |
| uptake of optimal doses of IPTp-SP implies uptake of at least three doses of IPTp-SP;<br>ANC – Antenatal care<br><sup>a</sup>p-values – overall p-values for each exposure variable in the model, CI – Confidence Intervals<br>OR – Odds ratios, crude OR (from Bivariate analysis), Adjusted OR (from Multivariable analysis)<br>**Goodness-of-fit of the model = F(9, 1262) = 0.222; p = 0.991**<br>**1 => Reference category** |  |  |  |  |  |

## Discussion

This study established the prevalence of IPTp-SP and the factors associated with the uptake of at least one dose and at least three doses of IPTp-SP among pregnant women in Nigeria. The study found a low and regional disparities in prevalence of IPTp-SP uptake during pregnancy. Several maternal characteristics such as, age group, education, antenatal care attendance, household wealth index and spouse’s educational attainment were found to contribute to the low IPTp-SP uptake during pregnancy in Nigeria.

In this study, compared with at least one SP dose at 63.6%, the prevalence of three or more SP doses was much lower at 16.8% among pregnant women in Nigeria. These findings fall significantly below the National Malaria Elimination Programme’s stipulated target for 100% of pregnant women attending ANC in Nigeria to receive at least three IPTp-SP doses by 2020 (24). A similar low prevalence of at least one dose at 62% and three or more IPTp-SP doses at 32% across sub-Saharan African countries were reported in the WHO malaria report in 2019 (1). According Hill *et al.,* 2013, differences in maternal characteristics across and within countries in sub-Saharan Africa contributed to low IPTp-SP prevalence of IPTp-SP (8). Also, Pons-Duran *et al.,* 2020 reported that the prevalence of taking optimal IPTp-SP doses was significantly below 25% in DR Congo, Madagascar and Nigeria (10). Conversely, the literature revealed the prevalence of IPTp-SP uptake is somewhat higher at 46%, 63% and 42% in Uganda (25), Ghana (13), and Malawi (26), respectively.

Several sociodemographic characteristics were established as factors associated with the uptake of at least one IPTp-SP dose. In terms of age group, women aged 35 years and above had higher odds of receiving at least one IPTp-SP dose during pregnancy. This finding was consistent with Olugbade *et al.,* 2019 in Nigeria (27) and Kibusi et al., 2015 in Tanzania (28). The higher uptake of at least one dose among older women may be attributed to earlier exposure to the benefit of IPTp-SP during prior pregnancies (29). In contrast, pregnant women’s age group did not play a significant role in taking optimal antimalarial regimens, as supported by Hill *et al.,* 2013 (8). Women residing in Northern Nigeria had the highest likelihood of taking at least one IPTp-SP dose due to a noticeable influence of Non-governmental Organisations in supporting increase access to healthcare interventions in the region (30). This regional variation in the level of IPTp-SP is consistent in other sub-Saharan African countries (8). This evidence may be due to inequality in accessing care in Nigeria, as Olukoya and Adebiyi reported in 2017 (23).

The finding agrees with Yaya et al., 2018, who found that pregnant women’s educational status in malaria-endemic countries negatively predicted optimal IPTp-SP doses (12). The result suggests that level of education can contribute to poor adherence to receiving optimal antimalarial regimens during pregnancy. The opposite is accurate regarding the association between uptake of IPTp-SP and the spouse’s educational level. The current findings agree with studies conducted in Eastern Nigeria (31). The findings infer that male involvement in ANC services during pregnancy might predict a higher IPTp-SP uptake (32). Utilising at least one dose and optimal IPTp-SP doses are associated with the household wealth index in different directions. Previous studies established a positive association between receiving optimal IPTp-SP doses and the household’s wealth index (10, 11). However, this current study found that pregnant women have reduced odds of receiving the optimal doses of IPTp-SP regardless of their wealth index. Conversely, Muhammed et al., 2020 found that pregnant women from the wealthiest households have increased IPTp-SP uptake than those from the poorest households in Nigeria (11). The disparity in the results may be attributed to the different methodologies adopted in assessing the association between wealth index and uptake of IPTp-SP. The current study reported that 84.3% of pregnant women who did not receive the optimal doses of IPTp-SP were also not exposed to media messages. The results may be a plausible reason for the wealth index indirectly influencing the uptake of optimal doses of IPTp-SP in this study. Regardless of the wealth index, studies support the evidence that knowledge of malaria-related interventions may affect adherence to completing three or more IPTp-SP doses during pregnancy (8, 15)

Early commencement of ANC services (timing of ANC initiation) implied an increased likelihood of initiating the first dose and completing at least three IPTp-SP doses during pregnancy. This finding aligns with the new IPTp-SP policy, which recommends the uptake of the first IPTp-SP dose after the first trimester during pregnancy and then successive IPTp-SP doses received in each scheduled ANC visit at a one-month interval (5). Studies conducted in Tanzania (16), Uganda (25), Malawi (33), and Nigeria (11) established similar findings. Also, ANC attendance was significantly associated with IPTp-SP uptake, as supported by Hill et al., 2013, a study conducted in SSA countries, including Nigeria (8). Among the study participants who visited the ANC clinic at least four times during pregnancy in 2018, approximately 84% of them initiated ANC services in their second trimester. Yet, only 23.3% received three or more IPTp-SP doses, though 80.3% received at least the first SP dose during pregnancy. This evidence implies lost opportunities to deliver the optimal IPTp-SP doses to pregnant women who visited ANC clinics at least four times. Even though the WHO recommends delivering IPTp-SP via scheduled ANC services to pregnant women living in malaria-endemic countries, the findings in Nigeria suggest otherwise. As evident in previous studies, the low IPTp-SP uptake via the ANC platform may be attributed to identified healthcare system constraints such as non-compliance to IPTp-SP guidelines (34), a shortage of trained healthcare workers (35), occasional SP stock-out (34, 35) and inadequate knowledge or awareness about IPTp-SP guidelines (36).

The finding suggests that exposure to malaria-related messages at least once a week may increase the odds of being aware of the benefits of IPTp-SP during pregnancy. This increased awareness may translate to higher chances of taking three or more IPTp-SP doses by pregnant women. Similar results have been found in Ghana (13), Uganda (25) and Tanzania (16). Furthermore, among pregnant, there was a direct association between taking IPTp-SP and their belief in its effectiveness against malaria infection. Such that their level of belief may explain their level of trust in the efficacy and safety of IPTp-SP. In this study, the findings suggest that women with less belief in the effectiveness of IPTp-SP were less likely to initiate IPTp-SP use than those with firm belief. This finding aligns with Balami et al., 2020, that knowledge about the efficacy of SP predicts the receipt of the first SP dose. And ultimately, the subsequent IPTp-SP doses during pregnancy (37). Also, pregnant women’s awareness of the morbidity caused by malaria predicted higher uptake of at least one SP dose but not optimal doses received. In agreement with the finding, Arnaldo *et al.,* 2019 found that a lack of awareness of malaria-associated morbidity reduced the odds of taking SP by pregnant women in Mozambique (38). These findings reiterate the need to utilise social and behaviour change communication strategies to drive IPTp-SP uptake at the community level. Meanwhile, there was no significant association among women who subscribed to health insurance and IPTp-SP uptake during pregnancy. In contrast, Darteh *et al.,* 2020 found an increased IPTp-SP uptake among women with health insurance coverage in Ghana (13). The difference in findings may be because only 2% of study participants were covered by health insurance in Nigeria compared to the study in Ghana where about 48% of study participants were covered by health insurance (13).

The major strength of this study was using a nationally representative sample to establish the factors related to IPTp-SP uptake. Next, the sample used was considerably large (n=12,742). Therefore, the findings may be generalisable to Nigerian women with live births on the factors that may influence the effective delivery of IPTp-SP during pregnancy. However, it is essential to note the following limitations in considering the factors associated with IPTp-SP use among women with live births in Nigeria. First, the established factors associated with IPTp-SP use do not imply causality because of the cross-sectional nature of the study design. Secondly, this study could not assess pregnant women’s attitudes and healthcare-related constraints concerning IPTp-SP use because the analysis was limited to the variables in the 2018 NDHS questionnaire. Therefore, there is a likelihood that women who attended at least four ANC visits during pregnancy did not receive IPTp-SP because of a stock-out of the drug. Third, excluding women who had stillbirths may have resulted in selection bias. The exclusion might have reduced the estimated effect of each possible factor on the uptake of IPTp-SP (39). Fourth, as a self-reported survey, the study is liable to social desirability bias as the women’s self-reported responses might not reflect the reality of the issues. Lastly, recall bias is an inherent limitation of survey designs resulting in varying degrees of accuracy in their previous experiences. However, some recall biases were minimised by including only women with live births during or two years before the Nigeria Demographic Health Survey in 2018.

## Conclusion

There is low uptake of at least one IPTp-SP dose and even lower uptake of three or more doses by pregnant women in Nigeria. This low uptake of IPTp-SP was associated with several maternal characteristics, age group, education, antenatal care attendance, household wealth index and trust in the efficacy and safety of SP. Therefore, there is a need for context-specific strategies such as targeted mass sensitisation and community awareness to increase the coverage of IPTp-SP uptake among vulnerable women. In addition, future research should explore the drivers of region-specific low uptake of optimal doses among pregnant women, especially in South-West Nigeria. IPTp-SP uptake remains suboptimal despite a relatively high antenatal care attendance among pregnant women in Nigeria. This situation calls for urgent action to deploy region-specific strategies to mitigate the bottlenecks (such as drug stock-out and non-compliance to the guidelines) at the healthcare facilities that hinder the delivery of IPTp-SP via ANC clinics in Nigeria. Also, pregnant women’s belief in the effectiveness of IPTp-SP influences their decision to initiate antimalarial treatment. Therefore, future research should examine the effectiveness of social-behavioural change communication strategies in driving uptake of three or more IPTp-SP doses during pregnancy in Nigeria.

## Data Availability

The dataset for the study was obtained with permission from the Demographic and Health Surveys (DHS) Program. The dataset are publicly available and may be requested from the DHS Program office via https://www.dhsprogram.com/data/dataset_admin/. The questionnaire used for the analyses is the women’s questionnaire contained within the 2018 NDHS.

https://www.dhsprogram.com/publications/publication-fr359-dhs-final-reports.cfm

## Acknowledgement

We are grateful to Measure DHS for the permission and for providing me with the dataset for this study. The authors also would like to thank the University of the Witwatersrand Human Research Ethics Committee (Medical) for the ethical approval.

## Authors contribution

**Conceptualisation**: Godwin Okeke Kalu, Joel Msafiri Francis, Juliana Kagura, Latifat Ibisomi

**Data curation**: Godwin Okeke Kalu

**Formal analysis**: Godwin Okeke Kalu, Joel Msafiri Francis, Juliana Kagura

**Methodology**: Godwin Okeke Kalu, Joel Msafiri Francis, Juliana Kagura, Latifat Ibisomi, Tobias Chirwa

**Funding Acquisition**: Latifat Ibisomi, Tobias Chirwa

**Project administration**: Godwin Okeke Kalu, Joel Msafiri Francis, Juliana Kagura

**Supervision**: Joel Msafiri Francis, Juliana Kagura

**Validation**: Godwin Okeke Kalu, Joel Msafiri Francis, Juliana Kagura

**Visualisation**: Godwin Okeke Kalu, Joel Msafiri Francis, Juliana Kagura

**Writing-original draft preparation**: Godwin Okeke Kalu

**Writing-review and editing**: Godwin Okeke Kalu, Joel Msafiri Francis, Juliana Kagura, Latifat Ibisomi, Tobias Chirwa

## Funding

TDR supported this research work as part of the Special Programme for Research and Training in Tropical Diseases (TDR). TDR is hosted by the World Health Organization and co-sponsored by UNICEF, UNDP, the World Bank and WHO. The TDR grant number: B40299. The funders did not play a role in the study design, data collection and analysis, decision to publish or preparation of the manuscript.

## Competing interests

The authors have declared no competing interests exist.

## Availability of data and materials

The dataset for the study was obtained with permission from the Demographic and Health Surveys (DHS) Program. The dataset are publicly available and may be requested from the DHS Program office via https://www.dhsprogram.com/data/dataset_admin/. The questionnaire used for the analyses is the women’s questionnaire contained within the 2018 NDHS, which can be accessed via https://www.dhsprogram.com/publications/publication-FR359-DHS-FinalReports.cfm.

## Supporting information

**S1 Table**. **Interaction term between Household wealth Index and Spouse’s Educational level.** This is the extension of Table 4 with interaction term

**S2 Table. Full multivariable logistic regression for the uptake of at least one IPTp-SP dose (Table 4)**. This is the STATA output for the final model (Table 4)

**S3 Table. Full multivariable logistic regression for uptake of at least three IPTp-SP doses**. This is the STATA output for the final model (Table 5)

**S4 Table. The model without interaction term for the uptake of at least one IPTp-SP dose**. This is the initial model without the interaction for factors associated with at least one SP dose.

**S5 Table. The model without interaction term for the uptake of at least one IPTp-SP dose**. This is the STATA output for the initial model without interaction term

## Notes

### Author Declarations

The Nigeria DHS 2018 was conducted by the Nigerian Population Commission in collaboration with the National Malaria Elimination Programme (NMEP) of the Federal Ministry of Health, Nigeria. Before each interview during the 2018 NDHS survey, all respondents provided informed consent, and the fieldworker ensured confidentiality. The respondents’ records were coded and de-identified (19). For this study, permission for NDHS 2018 dataset for secondary data analysis has been obtained from ICF International – Measure DHS website. Ethics approval was obtained from the University of the Witwatersrand Human Research Ethics Committee (Medical) – M2011103.

